# Right Or Left: Laterality Incidence Of Billed Plantar Fasciitis Plus Foot Pain

**DOI:** 10.1101/2022.06.09.22276198

**Authors:** Harry A. Kezelian, Ali A. Qadri, Shushovan Chakrabortty, Deepak Gupta

## Abstract

**Background:** Footedness and its potential role still remains largely underexplored in terms of laterality of lower extremity diseases.

**Materials and Methods:** Podiatric medicine patients’ electronic health/medical records were explored based on International Classification of Diseases, Tenth Revision, Clinical Modification (ICD-10-CM) codes billed from October 1, 2015-September 30, 2021 for pain in right foot (ICD-10-CM Diagnosis Code M79.671) as well as for plantar fasciitis (ICD-10-CM Diagnosis Code M72.2) vs. for pain in left foot (ICD-10-CM Diagnosis Code M79.672) as well as for plantar fasciitis (ICD-10-CM Diagnosis Code M72.2).

**Results:** During the six-year period, 1077 patients were only billed for pain in right foot vs. 921 patients were only billed for pain in left foot (p=0.0005) but only 362 patients with pain in right foot were concurrently billed for plantar fasciitis while only 372 patients with pain in left foot were concurrently billed for plantar fasciitis (p=0.71).

**Conclusion:** Although there was no significance difference between right vs. left plantar fasciitis incidence, the incidental finding of significant difference between right vs. left foot pain incidence warrants future investigations.

## Introduction

The potential role of footedness in relation to incidence of lower extremity diseases was recently explored in terms of lower extremity arthroplasty’s incidence [1]. However, footedness and its potential role still remains largely underexplored despite enormous databases of orthopedic and podiatric medicine patients [2] awaiting clinical researchers to ask the right questions about the laterality of their lower extremity diseases warranting medical interventions and/or surgical procedures therein.

## Materials and Methods

As institutional review board approved exempt research, podiatric medicine patients’ electronic health/medical records’ (EHR/EMR) database (NextGen Office, NextGen Healthcare, Inc, Atlanta, GA) at private clinic of author (HAK) were explored based on International Classification of Diseases, Tenth Revision, Clinical Modification (ICD-10-CM) codes billed from October 1, 2015-September 30, 2021 (six-year period). The total numbers of unique patient visits were quantified in terms of whether patients were billed for pain in right foot (ICD-10-CM Diagnosis Code M79.671) as well as for plantar fasciitis (ICD-10-CM Diagnosis Code M72.2) vs. whether patients were billed for pain in left foot (ICD-10-CM Diagnosis Code M79.672) as well as for plantar fasciitis (ICD-10-CM Diagnosis Code M72.2). These patient visits were also quantified and compared in terms of patients’ age and gender.

## Results

During the six-year period, 3629 patients were billed for pain in right foot (ICD-10-CM Diagnosis Code M79.671) and 3473 patients were billed for pain in left foot (ICD-10-CM Diagnosis Code M79.672) while 3432 patients were billed for plantar fasciitis (ICD-10-CM Diagnosis Code M72.2). However, on further analysis, it was determined that 2552 patients were billed for pain in right foot as well as pain in left foot so they were removed from the analysis as it could not be ascertained which sided pain might be related to plantar fasciitis. Among remaining 1077 patients who were only billed for pain in right foot vs. remaining 921 patients who were only billed for pain in left foot (p=0.0005), only 362 patients with pain in right foot were concurrently billed for plantar fasciitis while only 372 patients with pain in left foot were concurrently billed for plantar fasciitis (p=0.71). Demographic differences were also not statistically significant between these 362 right plantar fasciitis patients vs. 372 left plantar fasciitis patients (Table 1).

**Table 1:** Incidence Of Laterality In Plantar Fasciitis Per Billed Unique Patient Visits

|  | Right Plantar Fasciitis | Left Plantar Fasciitis | P-Value |
| --- | --- | --- | --- |
| Patients' Number (n) | 362 | 372 | 0.71 |
| Patients' Age (years)<br>(Mean ±SD) | 57.8 ±13.1 | 57.7 ±13.8 | 0.92 |
| Patients' Gender<br>(Females %) | 69% | 65% | 0.24 |

## Discussion

Contrary to our expectation of discovering right plantar fasciitis being more common than left plantar fasciitis due to right footedness’s preponderance in general population as well as right foot movements’ preponderance during automatic transmission vehicular driving [3-7], the podiatric medicine patients at private clinic of author (HAK) had insignificantly different incidence between right plantar fasciitis and left plantar fasciitis even though right foot pain incidence was significantly more common than left foot pain as per billed ICD-10-CM Diagnosis Codes’ data. This raises the question if planned future prospective clinical trials to ascertain incidence of laterality among plantar fasciitis patients at the author’s clinic will validate these findings or refute them.

There were some study limitations. Due to EHR/EMR inability to retrieve multiple visits by unique patients, it was not possible to discover any differences in frequency of follow-up visits between right plantar fasciitis patients and left plantar fasciitis patients. Moreover, as there is no separate ICD-10-CM Diagnosis Codes for right plantar fasciitis or left plantar fasciitis or bilateral plantar fasciitis, the billed data retrieved was an indirect analysis to compare the incidence of right plantar fasciitis vs. the incidence of left plantar fasciitis. Additionally, 2552 patients with billing for pain in right foot as well as pain in left foot (including 1508 patients with billing for plantar fasciitis as well) who were deleted from the final analysis might have validated or refuted the current results only if there were separate ICD-10-CM Diagnosis Codes for right plantar fasciitis and left plantar fasciitis. Moreover, 1190 patients who were billed for plantar fasciitis had no concurrent billing for pain in right foot or pain in left foot and hence could not be included in the final analysis due to missing data about laterality. Future retrospective and prospective studies by authors or global readers may be able to address and overcome these limitations.

## Conclusion

Although there was no significance difference between right vs. left plantar fasciitis incidence, the incidental finding of significant difference between right vs. left foot pain incidence warrants future investigations to refute or validate the hypothesis that predominance of right footedness and right foot based universal driving may be the underlying mechanism if right foot pain and/or right plantar fasciitis are found to be universally more common by researchers globally.

## Supporting information

IRB Approval

## Data Availability

All data produced in the present study are available upon reasonable request to the authors

